# Occurrence of cardiovascular complications associated with SARS-CoV-2 infection: a systematic review

**DOI:** 10.1101/2020.11.14.20231803

**Authors:** Daniele Melo Sardinha, Karla VB Lima, Thalyta MRL Ueno, Yan Correa Rodrigues, Juliana CD Garcez, Anderson LS Santos, Ana LS Ferreira, Ricardo JPS Guimaraes, Luana NGC Lima

## Abstract

Cardiovascular Diseases represent the main cause of death in the world, and are associated with risk factors that cause serious complications in cases of infections, such as those of the respiratory tract. In March 2020 the World Health Organization declared a pandemic for SARS-CoV-2, a new coronavirus causing severe pneumonia, which emerged in December 2019 in Wuhan, China. The objective is to investigate the occurrence of cardiovascular complications associated with SARS-CoV-2 infection. It is a systematic review, quantitative, in the databases, PubMed and Science direct, including primary studies with hospitalized patients confirmed for COVID-19 and who presented cardiovascular complications, the form used tools for evaluation of quality and evidence, following the PRISMA recommendations. Results: 12 studies were included. The occurrence of cardiovascular complications was: 27.35% of the sample of 3,316 patients. Types: Acute cardiac injury 17.09%; Thromboembolism 4.73%; Heart failure 3.43%; Arrhythmias 1.77%; stroke 0.33%. Mean age 61 years. Conclusions: This study showed that there are several cardiovascular complications associated with SARS-CoV-2, that the main one is acute cardiac injury, which causes several instabilities in the cardiopulmonary system, and that it is associated with mortality.

## 1. Introduction

Cardiovascular Diseases (CD) are a set of pathological conditions involving the cardiovascular system, such as Coronary Arterial Disease (CAD), Cardiac Arrhythmias, Valvopathy, Inflammatory Heart Disease, Heart Failure (HF), Vascular Brain Injury (VBI), Pulmonary Embolism (PE), and Deep Venous Thrombosis (DVT). However, some risk factors, modifiable and non-modifiable, potentiate or cause cardiovascular events, among them: Systemic Arterial Hypertension (SAH), Diabetes Mellitus (DM), Dyslipidemias, Sedentarism, Obesity, Stress, Hormone replacement therapy, Smoking, Family history of CD, Sex, Age and Heredity**[1,2]**.

Thus, CD represents a major public health problem, since it is the leading cause of death, accounting for 17.7 million deaths worldwide in 2015, representing 31% of all deaths globally. The main cause of death is CAD [3,4].

In this context, previous research has shown a transient risk of acute vascular events, including myocardial infarction and cardiovascular deaths, after clinically diagnosed Acute Respiratory Infections (ARI). Influenza vaccine has been shown to minimize the risk of major adverse cardiac events among people with pre-existing cardiovascular disease. Studies have shown that influenza epidemics are associated with cardiovascular mortality in subtropical and tropical climates. Thus, seasonal mortality from all causes, specifically in the elderly, has been associated with several viruses, including influenza, parainfluenza, RSV, and norovirus. A Dutch study suggests that these viruses account for 6.8% of deaths in people aged ≥85, 4.4% in people aged 75-84, and 1.4% in people aged 65-74. The impact of viral respiratory infections on the cardiovascular system causing fatal events is thus evident[5,6].

In this sense, in December 2019, cases of influenza syndrome with evolution to ARI, of unknown etiology, were registered in China and caused concern because of the several deaths and rapid transmissibility. In January researchers isolated and identified the causative infectious agent as a Coronavirus, named at the time 2019-nCov, however, due to the similarity to SARS-CoV, another coronavirus causing ARI in humans, was renamed by the World Health Organization (WHO) as SARS-CoV-2, and respectively the disease was named Cov Disease-19 (COVID-19)[7–9].

Soon, COVID-19, due to the high transmissibility via respiratory droplets, directly and indirectly, spread to neighboring countries and reached the level of the pandemic in March 2020 declared by the WHO[10].

According to the studies, the clinical characteristics of COVID-19 have been shown to evolve in three ways: Flu Syndrome (FS), Severe Acute Respiratory Syndrome (SARS), and Asymptomatic. In the FS it refers to the presentation of fever, cough, sore throat, coryza, nasal congestion, and myalgia. In SARS, signs, and symptoms of FS are associated with oxygen saturation equal to or <95%, beating of nose wings, diarrhea, nausea, vomiting, and cyanosis. The cases of SARS represent the serious complications of COVID-19, requiring intensive care treatment and being responsible for several deaths. The following risk factors stand out for the evolution of SARS: chronic heart diseases, chronic respiratory diseases, neoplasms, elderly, pregnant women, obese people[11–13].

In a review study, several cardiovascular complications associated with SARS-CoV-2 infection were cited[14]. However, studies do not specify the most frequent and prevalent associated complications, so it was questioned: What is the occurrence of cardiovascular complications associated with SARS-CoV-2 infection? The objective of this research is to investigate the occurrence of cardiovascular complications associated with SARS-CoV-2 infection.

## 2. Materials and Methods

Exploratory-descriptive research, of the type Systematic Review (SR), following the PRISMA recommendations [15]. The SR is made up of seven stages: 1. formulating a research question; 2. defining inclusion and exclusion criteria; 3. developing a research strategy and researching the literature - finding the studies; 4. selecting the studies; 5. evaluating the quality of the studies; 6. extracting the data; 7. summarizing the data and evaluating the quality of the evidence;[16].

For the elaboration of the research question, we used the PICO strategy, widely used in evidence-based practice, in which it proposes that problems identified in clinical practice, research, and teaching be organized from four elements: Patient; Intervention; Comparison; Outcome (PICO). Because the construction from these elements provides greater scope for the resolution of the problem addressed [17].

Research question: What is the occurrence of cardiovascular complications associated with SARS-CoV-2 infection cited in the literature? Patient: Infected with SARS-CoV-2 of any age group, hospitalized / Intervention: identify the cardiovascular complications associated with SARS-CoV-2 / Comparison: Not applicable / Outcome: Describe the occurrence of cardiovascular complications associated with SARS-CoV-2 cited in the literature.

For the search strategy in the literature, the following keywords were listed: cardiovascular complications; COVID-19; SARS-CoV-2. The selected databases were: PubMed, Science direct, including studies of any language, published from 2019 until the date of the search 11/07/2020. Inclusion criteria: primary studies, which evaluated individuals infected by SARS-CoV-2 hospitalized and presented results on cardiovascular complications, of all age groups. Exclusion criteria: studies that do not specify the cardiovascular complications that occurred quantitively.

The PRISMA flowchart, a tool that is part of the PRISMA protocol, was used to visualize the search in the databases, showing how the final sample was reached, describing all the stages, and inclusion and exclusion [15].

According to the protocol, two reviewers researched the bases independently, and critically evaluated each study from exhaustive reading in two stages, sorting by titles and abstracts/complete text. They then met twice to discuss the evaluations and decided which studies were included in the research.

The U.S. National Institutes of Health’s Quality Assessment Tool for Observational Cohort and Cross-Sectional Studies was used to assess the quality of studies (https://www.nhlbi.nih.gov/health-topics/study-quality-assessment-tools) It consists of 14 questions that measure the representativeness, the type of sampling, the description and evaluation of the exposure, the participants’ overbite and the adjustments and qualifications of the confounding variables. The results were discussed qualitatively among the researchers, and the disparities were solved through the discussion.

In the evaluation of the quality of evidence, the Oxford Centre for Evidence-Based Medicine’s Quality Classification Scheme was used for studies and other evidence [18], which is based on levels 1 to 5:

1. Randomized clinical trial properly fed and conducted; systematic review with meta-analysis.
2. Well designed controlled trial without randomization; a comparative prospective cohort study.
3. Case-control studies; a retrospective cohort study.
4. Series of cases with or without intervention; transversal study.
5. Opinion of respected authorities; case reports.

When extracting the data, the researchers developed a form consisting of the following variables: authors and year of publication, method, quality of evidence, location, age, participants, gender, comorbidities, estimates of the occurrence of cardiovascular complications evidenced in those infected by SARS-CoV-2. For data analysis, we opted for descriptive statistics. The results were presented in tables.

## 3. Results

In PubMed, the initial search resulted in 294 articles, after the analysis by title and objective, 46 were selected for reading. 7 articles were included for the data analysis [19], [20], [21], [22], [23], [24], [25]. In Science direct, 795 articles were initially searched, applying the filter for primary articles only resulted in 257. 45 were selected, and after reading 5, [26], [27], [28], [29], [30]. In figure 1 is the flowchart of the search and table 1 the detailing of the variables extracted from the studies.

**Table 1.** Data extracted from the studies included in the review.

| First author and year | Method | Quality of evidence | Location/sample | Age | Sex | Comorbidities | The occurrence and cardiovascular complications |
| --- | --- | --- | --- | --- | --- | --- | --- |
| Li et al (2020). | A retrospective study with patients Hospitalized | 3 | China<br>548 | Average 60 years<br>(range 48-69) | Male 279 (50,9%)<br>Female 269 (49,1%) | Chronic obstructive pulmonary disease 17 (3.1%)<br>Asthma 5 (0.9%)<br>Tuberculosis (1.6%)<br>Diabetes 83 (15.1%)<br>Hypertension 166 (30.3%)<br>Coronary heart disease 34 (6.2%)<br>Hepatitis B 5 (0.9%)<br>Chronic kidney disease 10 (1.8%)<br>Tumor 24 (4.7%) | Cardiac injury 119 (21.7%).<br>Diffuse intravascular coagulation 42 (7.7%). |
| Wang et al (2020). | The retrospective cohort in one hospital, all confirmed cases of COVID-19 over 60 years of age | 3 | China<br>339 | Average 71 years (> 60) | Male 166 (49%)<br>Female 173 (51%) | Heart Disease 53 (15.7%)<br>Hypertension 138 (40.8%)<br>Diabetes 54 (16%)<br>Kidney Disease 13 (3.8%)<br>Chronic Liver Disease 2 (0.6%)<br>Pneumopathy 21 (6.2%)<br>Autoimmune disease 5 (1.5%). | Acute cardiac injury 70 (21.0%)<br>Arrhythmia 35 (10.4%)<br>Heart failure 58 (17.4%) |
| Inciardi et al (2020). | The retrospective cohort with hospitalized patients | 3 | Italy<br>99 | Average 67 | Male 80 (81%)<br>Female 19 (19%) | Hypertension 63 (64%)<br>Dyslipidemia 29 (30%)<br>Diabetes 30 (31%)<br>Heart failure 21 (21%)<br>Atrial fibrillation 19 (19%)<br>Coronary artery disease 16 (16%) | Venous thromboembolism 12 (12%)<br>Arterial thromboembolism 3 (3%) |
|  |  |  |  |  |  | Chronic obstructive pulmonary disease 9 (9%)<br>Chronic kidney disease 15 (15%) Cancer 17 (18%) |  |
| Shi et al<br>(2020a). | A retrospective study with hospitalized patients | 3 | China<br>671 | Median 63<br>(50–72) | Male 322 (48%)<br>Female 349 (52%) | Hypertension 199 (29.7%)<br>Diabetes 97 (14.5%)<br>Coronary heart disease 60 (8.9%)<br>Chronic kidney disease 28 (4.2%)<br>Chronic obstructive pulmonary disease 23 (3.4%)<br>Cancer 23 (3.4%)<br>Heart Failure 22 (3.3%)<br>Cerebrovascular disease 22 (3.3%)<br>Atrial fibrillation 7 (1.0%) | Myocardial injury 106 (15.8%)<br>Heart Failure 12 |
| Li et al<br>(2020). | A retrospective observational study of hospitalized patients | 3 | China<br>219 | Median 53 | Male 89 (39.7%)<br>Female 130 (60.3%) | Hypertension 55 (25.1%)<br>Diabetes 31 (14.2%)<br>Heart disease 17 (7.8%) | Stroke 11 (5%) |
| Shi et al<br>(2020b). | Cohort with hospitalized | 3 | China<br>416 | Median 64<br>(21-95) | Male 205 (49.3%)<br>Female 211 (50.7%) | Hypertension 127 (30.5%)<br>Diabetes 60 (14.4%)<br>Coronary heart disease 44 (10.6%)<br>Cerebrovascular disease 22 (5.3%)<br>Heart Failure 17 (4.1%)<br>Chronic renal failure 14 (3.4%) | Cardiac Injury 82 (19.7%). |
|  |  |  |  |  |  | Chronic obstructive pulmonary disease 12 (2.9%)<br>Cancer 9 (2.2%) |  |
| Guo et al<br>(2020). | Series of<br>hospitalized<br>cases | 3 | China<br>187 | Average 58<br>(14-66) | Male 91 (48.7%)<br>Female 96 (51.3%) | Hypertension 61 (32.6%)<br>Coronary Heart Disease 21 (11.2%)<br>Cardiomyopathy 8 (4.3%)<br>Diabetes 28 (15.0%)<br>Chronic obstructive pulmonary disease 4 (2.1%)<br>Malignancy 13 (7.0%)<br>Chronic kidney disease 6 (3.2%) | Myocardial injury 52 (27.8%)<br>Coagulopathy 42 (34.1%) |
| Yang et al<br>(2020). | An observational<br>study of<br>hospitalized<br>patients | 3 | China<br>200 | Average 55 | Male 98 (49%)<br>Female 102 (51%) | Hypertension 45 (22.5%)<br>Chronic lung disease 7 (3.5%)<br>Diabetes 21 (10.5%)<br>Chronic heart disease 11 (5.5%)<br>Chronic kidney disease 3 (1.5%)<br>Chronic liver disease 2 (1%) | Acute cardiac injury 20 (10%) |
| Zhou et al<br>(2020). | Retrospective<br>and multicenter<br>cohort study,<br>with hospitalized | 3 | China<br>191 | Average 56<br>(18-87) | Male 119 (62%)<br>Female 72 (38%) | Hypertension 58 (30%)<br>Diabetes 36 (19%)<br>Coronary heart disease 15 (8%)<br>Chronic obstructive pulmonary disease 6 (3%)<br>Carcinoma 2 (1%) | Heart failure 44 (23%)<br>Coagulopathy 37 (19%)<br>Acute cardiac injury 33 (17%) |
| Zhang et al<br>(2020). | A retrospective<br>study with<br>hospitalized | 3 | China<br>221 | Average 55<br>(39-66) | Male 108 (48.9%)<br>Female 113 (51.1%) | Hypertension 54 (24.4%)<br>Diabetes 22 (10.0%)<br>Cardiovascular disease 22 (10.0%)<br>Cerebrovascular disease 15 (6.8%) | Arrhythmia 24 (10.9%)<br>Acute cardiac injury 17 (7.7%) |
|  |  |  |  |  |  | Chronic obstructive pulmonary disease 6 (2.7%)<br>Chronic kidney disease 6 (2.7%)<br>Chronic liver disease 7 (3.2%)<br>Immunosuppression 3 (1.4%) |  |
| Shang et al<br>(2020). | Retrospective<br>cohort study with<br>severe<br>hospitalized | 3 | China<br>113 | Median 66 | Male 73 (64%)<br>Female 40 (36%) | Hypertension 50 (44%)<br>Diabetes 20 (17.7%)<br>Coronary heart disease 28 (24.8%)<br>Chronic obstructive pulmonary disease 5 (4%)<br>Tumor 8 (7.1%)<br>Heart disease 9 (8.0%)<br>Liver disease 8 (7.1%) | Acute myocardial injury 44 (38.9%)<br>Coagulopathy 21 (18.6%) |
| Deng et al<br>(2020). | Retrospective<br>with hospitalized | 3 | China<br>112 | Average 65<br>(49-60) | Male 57 (50.9%)<br>Female 55 (49.1%) | Chronic obstructive pulmonary disease 4 (3.6%)<br>Hypertension 36 (32.1%)<br>Diabetes 19 (17.0%)<br>Coronary artery disease 15 (13.4%)<br>Atrial fibrillation 4 (3.6%) | Myocardial injury 14 (12.5%) |

Source: Authors.

**Figure 1.**
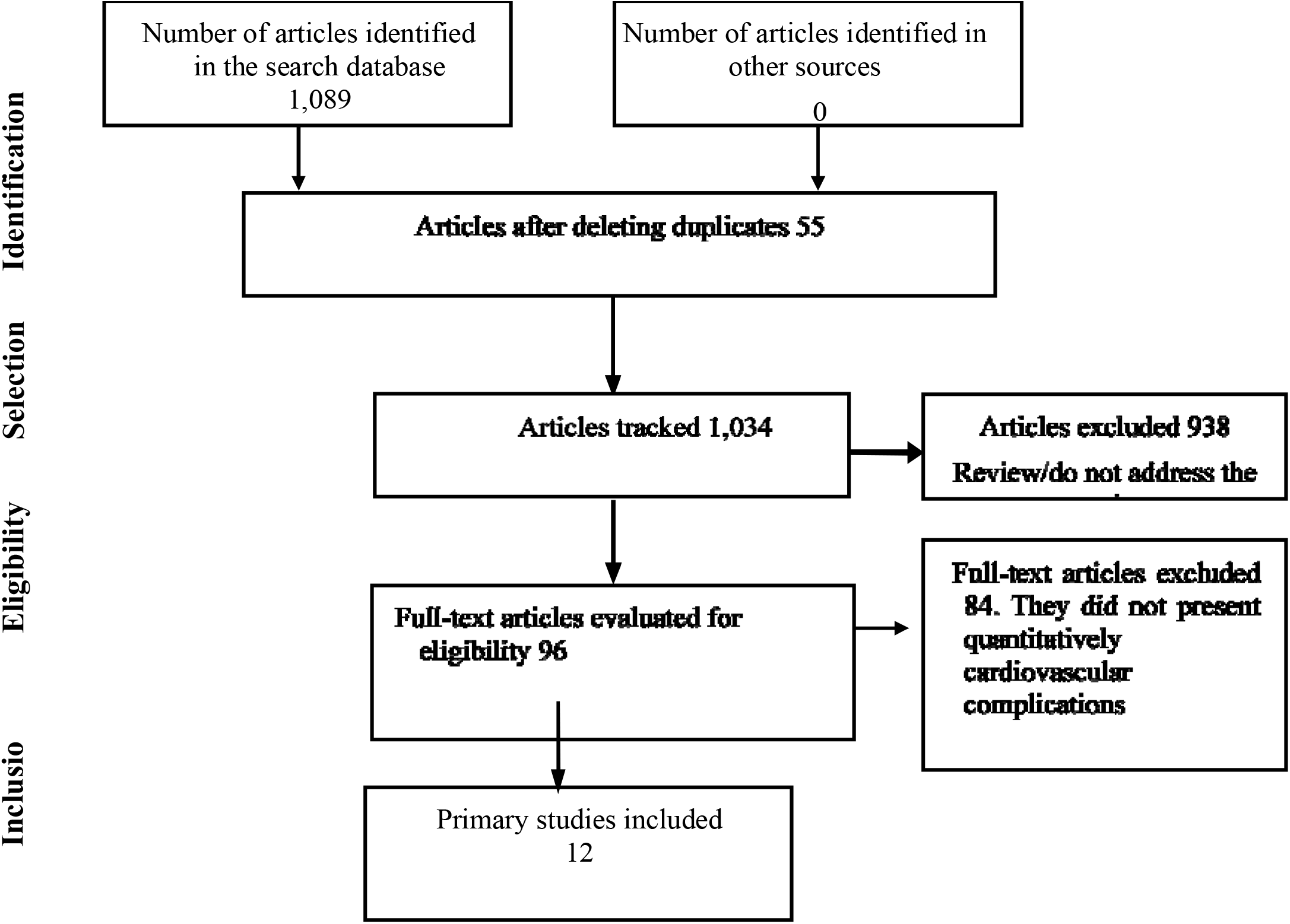
Characterization of the search and definition of the sample in the databases.

Based on the analysis of the included studies and the extracted variables, the review gathered 3. 316 participants of the studies who were hospitalized, 50.87% male and 49.12% female, mean age 61 years, 13.08% with heart diseases (heart failure and coronary disease were included in heart diseases), 15.10% diabetes, 31.72% hypertension, 3.5% pneumopathy (obstructive pulmonary disease was included in pneumopathy), 3.13% renal diseases, 2.89% cancer, 0. 09% immunosuppression, 0.57% liver disease, 0.15% autoimmune disease and 0.27% tuberculosis. This shows the most frequent comorbidities are: Hypertension, Diabetes, and Cardiac Diseases (table 2).

**Table 2.** Characteristics of participants, gender, mean age, and comorbidities.

|  | Yes |  | No |  | Total |  |
| --- | --- | --- | --- | --- | --- | --- |
|  | n | % | n | % | n | % |
| Female | 1629 | 49,12545 | 1687 | 50,87455 | 3316 | 100 |
| Male | 1687 | 50,87455 | 1629 | 49,12545 | 3316 | 100 |
| Age (average) 61 years |  |  |  |  | 3316 | 100 |
| Heart disease | 434 | 13,08806 | 2882 | 86,91194 | 3316 | 100 |
| Diabetes | 501 | 15,10856 | 2815 | 84,89144 | 3316 | 100 |
| Hypertension | 1052 | 31,72497 | 2264 | 68,27503 | 3316 | 100 |
| Pneumopathy | 119 | 3,588661 | 3197 | 96,41134 | 3316 | 100 |
| Chronic kidney disease | 104 | 3,136309 | 3212 | 96,86369 | 3316 | 100 |
| Cancer | 96 | 2,895054 | 3220 | 97,10495 | 3316 | 100 |
| Immunosuppression | 3 | 0,09047 | 3313 | 99,90953 | 3316 | 100 |
| Liver disease | 19 | 0,572979 | 3297 | 99,42702 | 3316 | 100 |
| Autoimmune disease | 5 | 0,150784 | 3311 | 99,84922 | 3316 | 100 |
| Tuberculosis | 9 | 0,271411 | 3307 | 99,72859 | 3316 | 100 |

Source: Authors.

Regarding the occurrence of cardiovascular complications associated with the SARS-CoV-2 infection, the following were highlighted: acute cardiac injury 17.09%, thromboembolism 4.73%, heart failure 3.43%, arrhythmias 1.77%, and stroke 0.33% (Table 3).

**Table 3.**
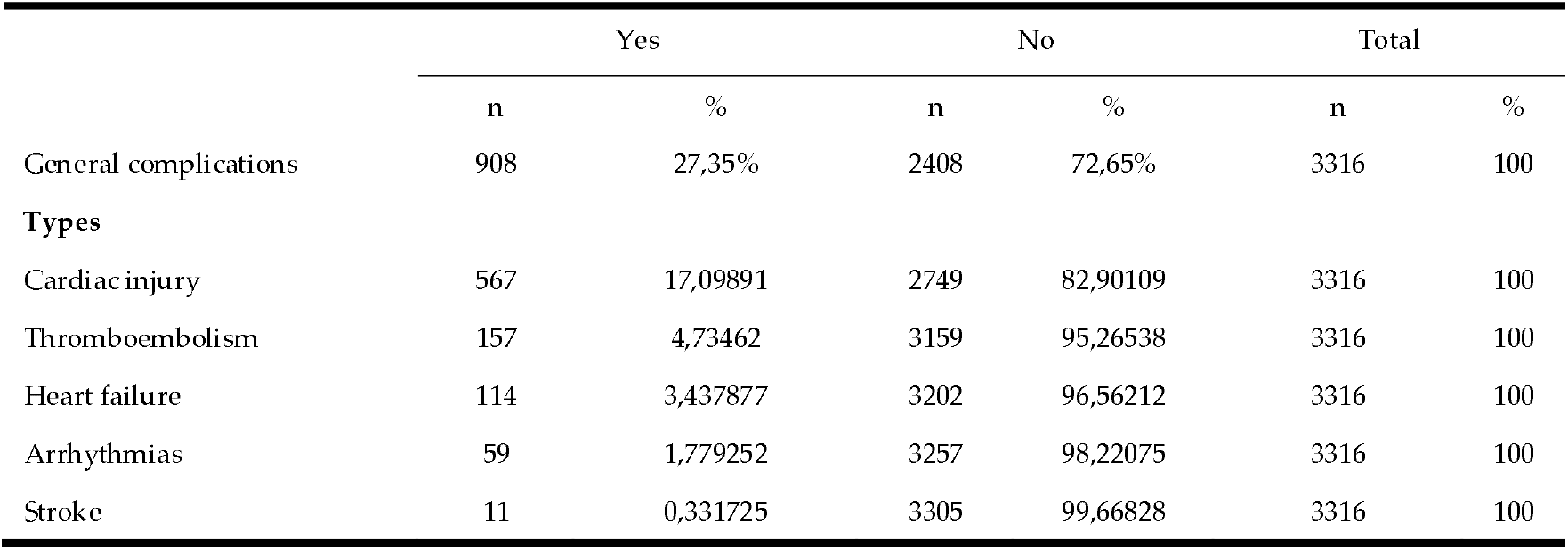
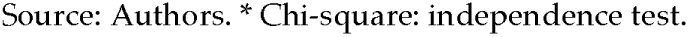
Occurrence of cardiovascular complications associated with SARS-CoV-2.

|  | Yes |  | No |  | Total |  |
| --- | --- | --- | --- | --- | --- | --- |
|  | n | % | n | % | n | % |
| General complications | 908 | 27,35% | 2408 | 72,65% | 3316 | 100 |
| **Types** |  |  |  |  |  |  |
| Cardiac injury | 567 | 17,09891 | 2749 | 82,90109 | 3316 | 100 |
| Thromboembolism | 157 | 4,73462 | 3159 | 95,26538 | 3316 | 100 |
| Heart failure | 114 | 3,437877 | 3202 | 96,56212 | 3316 | 100 |
| Arrhythmias | 59 | 1,779252 | 3257 | 98,22075 | 3316 | 100 |
| Stroke | 11 | 0,331725 | 3305 | 99,66828 | 3316 | 100 |

## 4. Discussion

Infection with SARS-CoV-2 may manifest with mild symptoms (85%) and severe symptoms (15)% (such as progression to SARS) with the potential to cause extrapulmonary damage, including cardiovascular damage that directly influences morbidity and mortality [31].

Thus, this study showed, from the analysis of studies with 3,316 hospitalized patients confirmed for COVID-19, that the occurrence of cardiovascular complications associated with SARS-CoV-2 was 27.35% of the following types: Acute cardiac injury 17.09%; Thromboembolism 4.73%; Heart failure 3.43%; Arrhythmias 1.77%; stroke 0,33%.

The acute cardiac injury was the most recurrent, and two meta-analysis studies showed its association with increased troponin and mortality, showing no association of cardiac injury with a higher risk of developing SARS. The studies highlight that early troponin monitoring is a preventive strategy for further myocardial injury[32,33]. This mechanism of injury is not yet clear to cardiologists, however, they work with hypotheses of viral invasion in the myocardium, similar to pulmonary invasion, via the connection of SARS-CoV-2 to the Angiotensin-Converting Enzyme 2 (ACE2), which is extensively present in the myocardium. Additionally, pneumonia can influence systemic inflammatory responses, causing non-ischemic complications in the myocardium, mainly in the presence of cardiovascular comorbidities. It is also noted that the increased inflammatory activity is a risk factor for the rupture of coronary atherosclerotic plaques, which can cause the partial or total blockage of coronary arteries, causing myocardial ischemia and consequently hypoxia injury, characterizing Acute Myocardial Infarction. Causes of cardiomyopathy due to stress, physical injury, or pharmacological effects were not excluded.[34,35].

Research characterizes that the occurrence of acute cardiac injury is highly related to the presence of cardiovascular comorbidities (such as hypertension and diabetes), admission to an Intensive Care Unit, and mortality. They highlight as a characteristic the increase of troponin type I associated with electrocardiographic and echocardiographic alterations, presenting segmental abnormality of the wall movement or reduction of the left ventricle ejection fraction [36]. Thus, the clinical presentation of the acute cardiac lesion may manifest as myocarditis or acute myocardial infarction, with thoracic pain and ST-segment elevation, requiring percutaneous cardiac procedure[37,38], which is an intervention that presents a risk of acute renal failure in hospitalized patients with comorbidities, due to the use of contrasts, and can increase hospitalization and mortality [39]. Showing that this complication requires procedures that can influence or enhance clinical conditions for a worse outcome.

Thromboembolism was the second most recurrent cardiovascular complication, described in the literature as Disseminated Intravascular Coagulation. In association with SARS-CoV-2, it is related to a storm of inflammatory cytokines, directly attacking the vascular endothelium with an elevation of the fibrinogen and D-dimer markers (product of fibrin/fibrinogen degradation), causing thrombin dysregulation, exacerbated by inhibition of fibrinolysis. Thus the natural anticoagulants are compromised, resulting in the formation of thrombi, which can travel to a smaller vessel and obstruct, causing hypoxia. Thromboses can be venous or arterial and can evolve to Deep Venous Thrombosis, Pulmonary Embolism, Ischemic Vascular Accident, or acute myocardial infarction. These are serious complications associated with mortality. In this context, the heparin used in anticoagulant therapy was associated with a better prognosis in some cases[40,41]. A study with 184 patients hospitalized in an intensive care unit confirmed for COVID-19, showed an incidence of 31% of thrombotic events, all received standard-dose tombroprophylaxis[42]. In another study with 388 patients hospitalized by COVID-19 with thrombotic events in 28 cases (7.7%), being 8 cases in the intensive care unit and 20 cases in the infirmary, the complications represented: 4.4% venous thromboembolism, 2.8% Pulmonary Embolism, 0.3% Deep Venous Thrombosis, 2.5% ischemic stroke and 11.1% acute myocardial infarction and Acute Coronary Syndrome. All cases used tombroprophylaxis[43]. This study also showed the occurrence of stroke alone as a complication in 0.33% of cases, however, it is directly associated with thromboembolism associated with SARS-CoV-2, because the inflammatory response generates a hypercoagulable condition.

Heart failure represented the third cardiovascular complication, being a complex clinical syndrome, in which the myocardium cannot adequately pump the blood to meet the metabolic needs of the tissues. This condition can be caused by structural or functional cardiac changes and presents characteristic signs and symptoms that result from a reduction in cardiac output and/or high filling pressures at rest or under stress[44]. Studies show that this complication occurs in patients without heart disease, mainly as a consequence of acute heart injury, since the damage to the myocardium has repercussions on its ventricular contraction function, resulting in a reduction in systolic function. Additionally, coagulopathies can cause pulmonary embolism and have repercussions in acute right ventricular failure. Stress cardiomyopathy can also lead to classic ventricular decompensation, with high filling pressures and pulmonary edema. On the other hand, in the carrier of chronic heart failure, there is a great chance of decompensation of the disease, requiring hospitalization and intensive care, influencing mortality[45,46].

Another outstanding complication was cardiac arrhythmias, which are directly related to the following factors: acute cardiac injury, heart failure, drugs that prolong the qt interval. Atrial fibrillation proved to be the most common type of arrhythmia, with the potential risk of thrombus formation, acute myocardial infarction, and ischemic stroke, and antithrombotic therapy is debatable even in cases without risk factors. Arrhythmias are also related to higher risks of heart instability, cardiac arrest, and death[47].

This review showed that acute cardiac injury is the main cardiovascular complication associated with SARS-CoV-2 infection, causing several instabilities in cardiopulmonary functions, and also influencing other cardiovascular complications, in patients without chronic diseases and with chronic diseases.

## 5. Conclusions

The main cardiovascular complications associated with SARS-CoV-2 in hospitalized patients were several, among them: acute cardiac injury, thromboembolism, heart failure, arrhythmias, and ischemic stroke, with a mean age of 61 years, with the most prevalent comorbidities, hypertension, diabetes, and heart diseases.

The evidence on the association of cardiovascular complications and SARS-CoV-2 provides information for the implementation of preventive measures and better therapeutic management, to minimize hospital stay, morbidity, and mortality.

## Data Availability

Estao disponiveis de forma publica

